# Multi-stain modelling of histopathology slides for breast cancer prognosis prediction

**DOI:** 10.1101/2024.11.10.24317066

**Authors:** Abhinav Sharma, Fredrik K. Gustafsson, Johan Hartman, Mattias Rantalainen

## Abstract

**Background:** Pathologic assessment of the established biomarkers using standard hematoxylin & eosin (H&E) and immunohistochemical (IHC) stained whole slide images (WSIs) is central in routine breast cancer diagnostics and contributes prognostic and predictive information that guides clinical decision-making. However, other than only aggregated protein-expression values from IHC WSIs, a spatial combination of histo-morphological information from IHC and H&E WSIs can potentially improve prognosis prediction in breast cancer patients. In this study, we aim to develop a deep learning-based risk-stratification method for breast cancer using routine H&E and IHC-stained histopathology WSIs from resected tumours.

**Methods:** This is a retrospective study including WSIs from surgical resected specimens from 945 patients from the South General Hospital in Stockholm. One H&E and four IHC (ER, PR, HER2, and Ki-67) stained sections were included from each patient, retrieved from the same tumour block. The IHC WSIs with the H&E WSI were registered, and corresponding images patches (tiles) were extracted for each image modality. Features from the registered tiles were extracted using two existing and publicly available histopathology foundation models (UNI and CONCH). Using the extracted features together with time-to-event data, we optimised an attention-based multiple instance learning (MIL) model using the Cox loss (negative partial log-likelihood loss) and recurrence-free survival (RFS) as the survival endpoint.

**Results:** Using cross-validation we observed a prognostic performance with a C-index of 0.65 (95%CI: 0.56 - 0.72) for the risk score prediction using only H&E WSIs and UNI as the tile-level feature extractor. Combinations of H&E with one or more IHC modalities were subsequently evaluated, with the highest performance observed in the model combining the H&E and PR WSI data and the model combining all the stains, obtaining a C-index of 0.72 (95% CI: 0.65 - 0.79) and 0.72 (95% CI: 0.64 - 0.79) respectively.

**Conclusion:** Multiple stain modalities are used in routine breast cancer pathology, but has not been considered together for prognostic modelling. The results in this study suggests that models combining morphological features extracted by histopathology foundation models across multiple stain modalities can improve prognostic risk-stratification performance compared to single-modality models.

## Introduction

Breast cancer histopathology specimens are routinely assessed using several staining, including general morphological features using the Hematoxylin and Eosin (H&E) stain, and specific biomarkers using Immunohistochemical (IHC) staining. Histological grading is a prognostic factor that is determined by assessing H&E slides, whereas assessment of biomarkers such as Estrogen receptor (ER), Progesterone receptor (PR), Human epidermal growth receptor-2 (HER2) and Ki-67 is performed using the respective IHC (ER, PR, HER2, and Ki-67) stained slides in the routine diagnostic workflow [1–4].

In recent years, the digitisation of routine pathology has enabled the development of deep learning-based models for high-resolution whole-slide images (WSIs). Various applications include biomarker prediction, gene expression prediction, and treatment response prediction among others that show the potential of such models to assist pathologists in diagnostics and clinical decision-making [5–11]. There have been studies reporting histology-based prognosis prediction from WSI that include patient risk-stratification, risk-of-recurrence prediction, and treatment response prediction [10,12–14]. The prognostic risk score prediction task involves the modelling of time-to-event from diagnosis to the observed outcome [15]. Recently, multi-modal prediction models reported the combination of different modalities such as histology, gene expression and clinical covariates [12,16–19]. They have shown to improve the risk score prediction performance over a single data modality.

In routine diagnostics, H&E has been one of the standard and most widely used stains for more than a century [20]. Given the high-resolution of histo-morphological information present in H&E WSIs, most of the histology-based modelling of prognostic risk score prediction included only H&E WSIs [14,21,22]. However, routine IHC stains also include prognostic information that could be utilised for modelling the patient risk-stratification. The IHC-based prognostic markers are assessed by the aggregated protein expression in IHC WSIs and used as the binary status for clinical decision-making [23]. The local and spatial presence of protein expression along with the information present in the counterstain in IHC, when combined with local histo-morphological information from other stain modalities can potentially incorporate more prognostic information that can further improve the prognosis prediction in breast cancer patients. Previously, multi-stain modelling of WSIs has been reported for colorectal cancer recurrence prediction models and shown to have improved the prediction performance with respect to single-stain WSI modalities [24]. To the best of our knowledge, we have not observed any studies reporting the multi-stain risk score prediction for breast cancer patients.

In this study, we aim to develop and validate the deep learning-based multi-stain modelling for risk score prediction in breast cancer patients using H&E and routine IHC WSIs. We include H&E WSIs with ER, PR, HER2 and Ki-67 WSIs for the joint model that has the potential to combine the spatial histo-morphological information between different stains which can further improve the prognostic performance in breast cancer patients.

## Methods

### Datasets

In this retrospective observational study, we used the SöS-BC-4 [25] cohort that includes invasive breast cancer patients from the South General Hospital (Södersjukhuset) in Stockholm, Sweden, diagnosed between 2012-2018. One H&E and four IHC stained WSIs from the same resected tumour block were used for each patient. The clinical variables for the patients were derived from the National Breast Cancer Registry (NKBC) in Sweden. Patients with missing clinical variables were excluded from the study. In total 945 patients were included in the study (Figure 1a).

**Figure 1:**
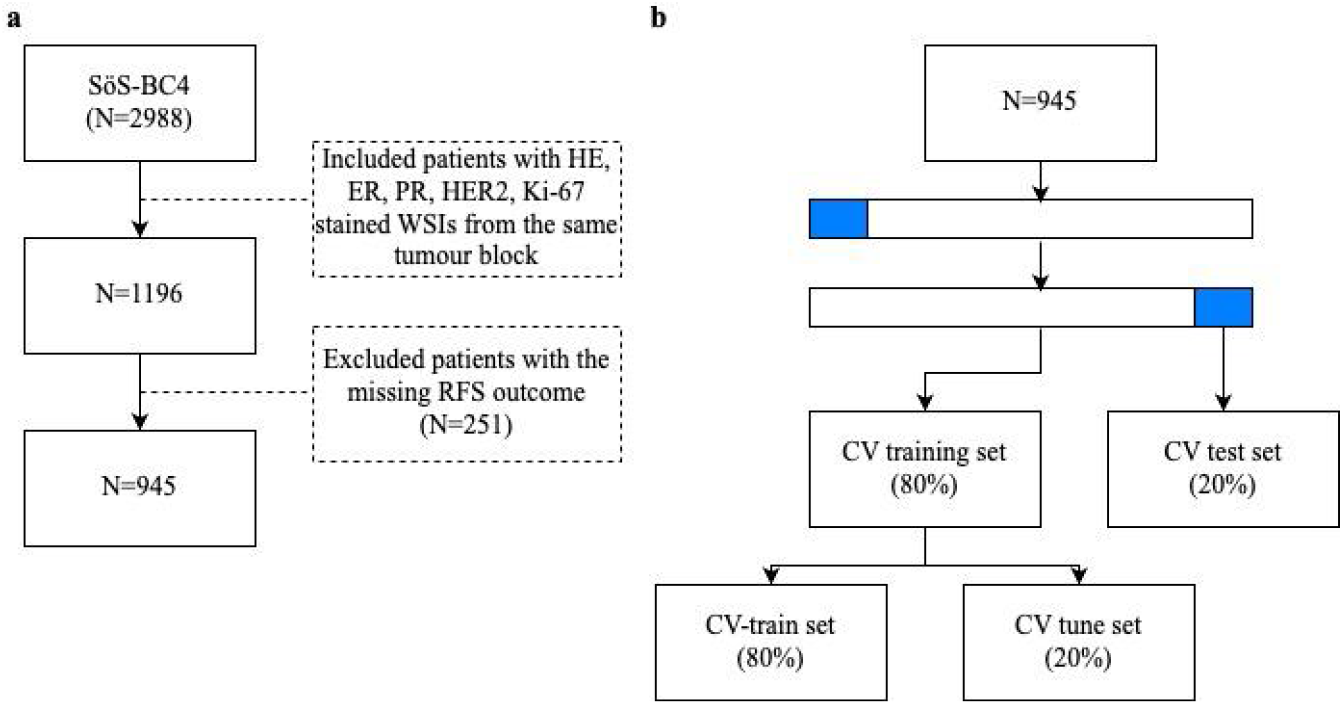
CONSORT diagram. **a** Inclusion and exclusion criterias for the SöS-BC4 cohort used in the study. We included the Hematoxylin & Eosin (H&E) whole-slide images (WSIs) from the patients with all four immunohistochemistry (IHC) WSIs from the same tumour block. Further, we excluded patients with missing recurrence-free survival (RFS) outcomes. **b** 5-fold stratified cross-validation (cv) splits were used to optimise the risk score prediction models.

In this study, the 5-fold cross-validation was performed using patient-level (WSI-level) stratified splits based on clinicopathological variables: Age, ER status, PR status, HER-2 status, Lymph node status and Tumour Size. The training set was further split into CV-train and CV-tune sets for model optimisation.

### WSI preprocessing

The detailed preprocessing steps for H&E WSIs are described in [10]. Firstly, it included the tissue masking step to remove the background from the WSI. Then, we tiled the WSI into tiles of size 1196×1196 pixels at 40x and downscale it by a factor of two to retrieve tiles of size 598×598 pixels at 20x resolution (0.45 microns per pixel). Next, the tiles with the variance of the Laplacian filter < 500 units were considered blurry and excluded from the further analyses. Lastly, we applied the colour normalisation method as described by [26] but with a slight variation to enable the WSI-level colour correction, as described in [10].

#### WSI registration

Then, we performed the registration of each IHC WSI modality with the corresponding H&E WSI using the non-rigid registration method as described in [27]. After preprocessing the H&E WSIs, we retrieved the coordinates of each H&E tile and transformed the centroid of the H&E tiles with the registered IHC modality as the landmarks. Further, we retrieved the centroid coordinates on the registered IHC WSI and extracted tiles of size 1196×1196 pixels at 40x resolution (as shown in Figure 2a).

**Figure 2.**
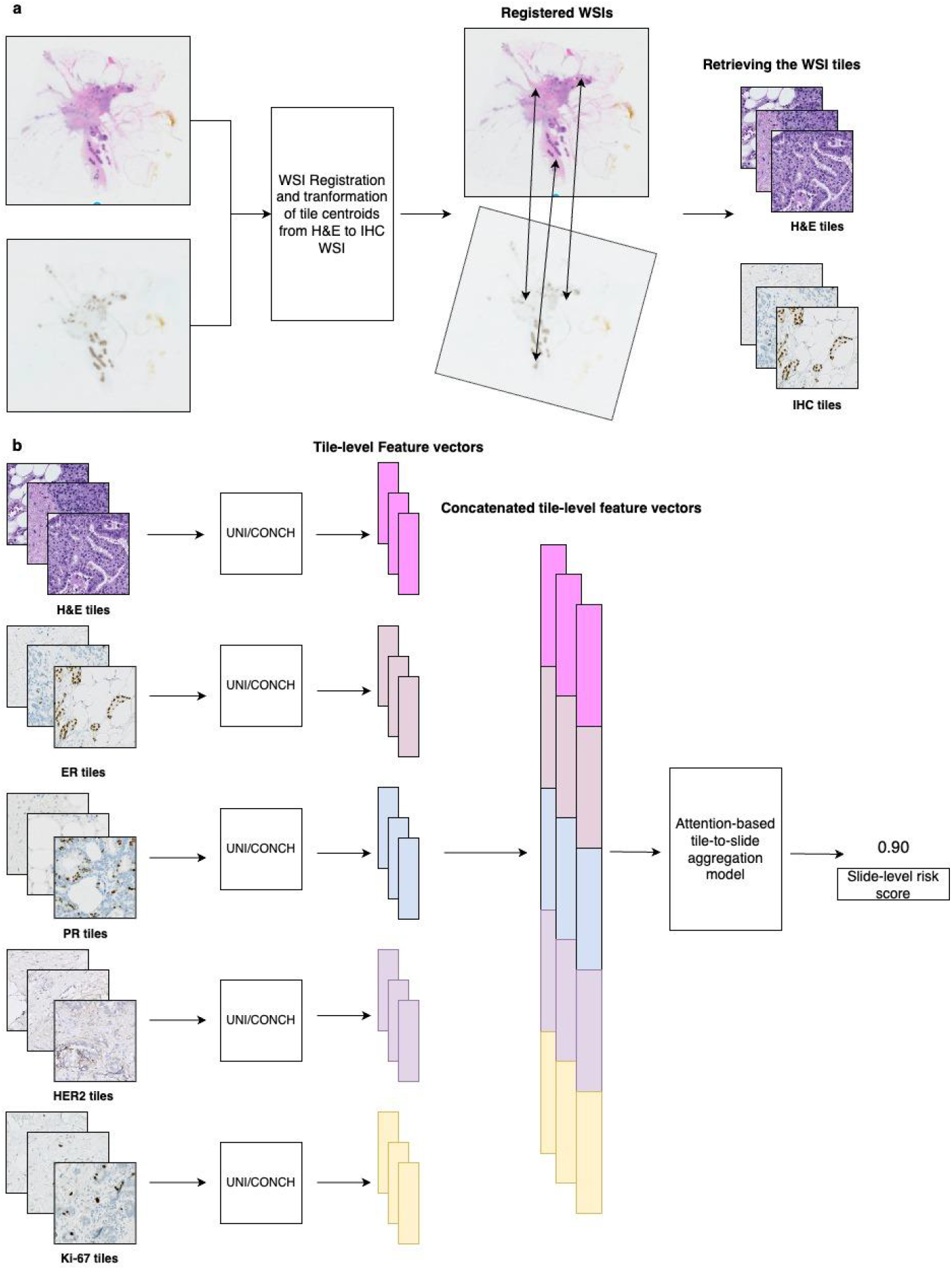
Overview of the study design. **a** First, the non-rigid registration method was applied to each IHC WSI with the corresponding H&E WSI. Tile centroid coordinates were transformed from the H&E WSI to the registered IHC. Then, using the corresponding tile centroids, tiles of the same size as the H&E tiles from the IHC WSI (598 × 598 pixels at 20x magnification) were created. **b** Next, feature vectors from each tile using the foundation models i.e. UNI and CONCH were extracted. Further, we concatenated the feature vectors from corresponding tiles from each stain modality and then optimised the tile-to-slide aggregation model (ABMIL) with concatenated features with Cox loss to provide the patient-level risk score.

### Modelling strategy

We extracted features from the tiles using two publicly available foundation models: UNI [28] and CONCH [29] (Figure 2b). UNI is a foundation model trained using a self-supervised learning method called DINOv2 [30] on 100k H&E WSIs across multiple tissue types [28]. We used the image encoder from CONCH, which is a vision-language foundation model trained across 1.1 million pathology image-caption pairs [29]. UNI is based on a ViT-L architecture that takes images of size 224×224 pixels and extracts feature vectors of dimension 1024, whereas the image encoder in CONCH is based on a ViT-B architecture that takes input images of size 448×448 pixels and extracts feature vectors of dimension 512. We applied the RandomCrop transform from the torchvision package [31] to transform the tile size to desired input sizes for UNI and CONCH models respectively.

After retrieving the frozen tile-level feature vectors for corresponding tiles from each stain modality, we first concatenated the feature vectors (Figure 2b). Then, we optimise an attention-based Multiple Instance Learning (ABMIL) model using the concatenated feature vectors, inspired from the implementation of the model CLAM [32] without the instance-level classifier. It aggregates the tile-level feature vectors to a slide-level feature vector and then the network head includes the regression layer that takes slide-level features as input and outputs a continuous slide-level risk score for each patient. The ABMIL model was optimised using the negative partial log-likelihood loss of the Cox Proportional Hazards model (i.e. Cox loss) with Recurrence-Free Survival (RFS) as the survival endpoint [33]. The RFS was defined as the recurrence (i.e. local or distant metastasis, detection of contralateral tumours) or death as the event outcome. Patients were followed from the initial time of diagnosis to the date of recurrence/death, emigration, or the last registration date, whichever occurred first. We created WSI bags with tile-level feature vectors and created batches of 24 bags for each step per epoch. We used Adam [34] as the optimiser with a momentum of 0.9.

### Statistical analyses

We retrieved the patient-level risk score for each CV test set from the optimised ABMIL models. Then, we aggregated the risk scores from all the CV test sets for the evaluation of predictive performance using the concordance index (C-index) as the performance metric.

## Results

Firstly, we evaluated the 5-fold CV risk-stratification performance with each individual stain modality (Figure 3a). We observed the C-index for risk score prediction using only the H&E WSI to be 0.65 (95%CI: 0.54 - 0.74) and 0.64 (95%CI: 0.56 - 0.72) with UNI and CONCH as the feature extractor, respectively. The C-index for risk score prediction with only PR WSIs was observed to be relatively higher than the prediction using other individual IHC stains (C-index: 0.62 (95% CI: 0.57 - 0.74)) using UNI as the feature extractor. We observed the point estimates for C-index to be higher for three out of five individual stain modalities using UNI in comparison to CONCH.

**Figure 3.**
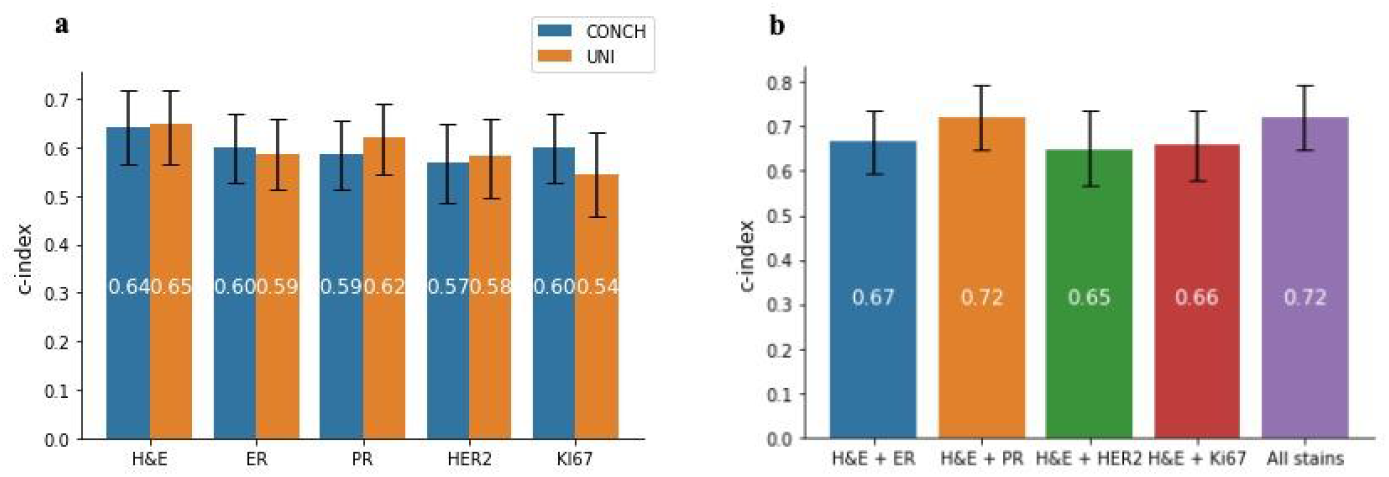
Risk score prediction performance with individual and multi-stain whole-slide image modelling approaches. **a** Concordance-index (C-index) for the risk score prediction using individual stain WSIs. For each stain modality, we used UNI and CONCH as tile-level feature extractors and optimised a ABMIL model for risk score prediction and reported the aggregated 5-fold cross-validation C-index. The error bar shows the lower and upper limits of the bootstrapped 95% confidence interval. **b** Risk score prediction using concatenated features from multiple stain WSIs. The frozen UNI tile-level feature vectors from the H&E stain were concatenated with tiles from each IHC stain modality. The last bar includes the concatenation of tile-level feature vectors from all five stain modalities.

Next, we used UNI as the tile-level feature extractor for all stain modalities. Then, we concatenated the frozen tile-level features from each stain modality to optimise the ABMIL model for risk score prediction using 5-fold CV. Initially, we observed the risk score prediction performance by concatenation of tile-level feature vectors from each stain modality to H&E, and then concatenation of tile-level features from all five stain modalities (Figure 3b). We observed a similar C-index between the concatenation of H&E with PR tile-level feature vectors (H&E + PR) and H&E with all the IHC tile-level feature vectors (C: 0.72).

Then, we compared prognostic risk score prediction performance using a model that only included clinical variables (Age, Tumour Size, Lymph Node status and histological grade). The patients with missing clinical covariates were excluded from the model (N=803). We observed a C-index of 0.67 (95% CI: 0.58 - 0.76) based on the Cox Proportional Hazard (PH) model fitted with clinical variables only using 5-fold CV (Figure 4). The multi-modal models were re-optimsed on the cohort subset (N=803) to enable direct comparison with clinical only model. Finally, we evaluated a model where we concatenated the clinical variables to the slide-level feature vectors from H&E + PR and All stains model, while optimising the ABMIL model. We observed marginal improvement in the risk score prediction performance by adding clinical variables to the H&E + PR model (C-index: 0.72 (95% CI: 0.63 - 0.79)), however did not observe improvement in All stains model 0.70 (95%CI: 0.60 - 0.78) (Figure 4).

**Figure 4.**
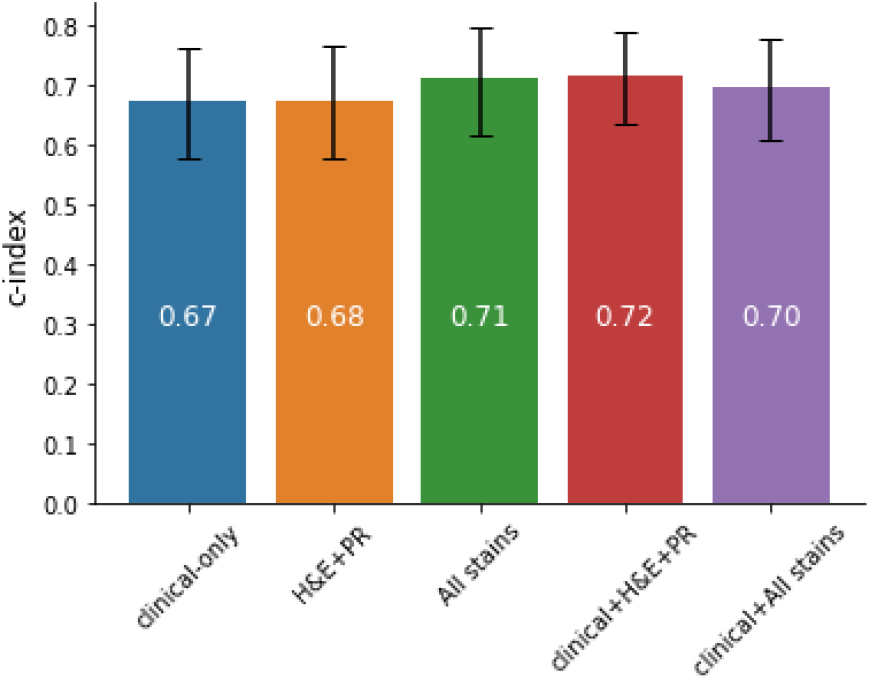
Risk score prediction performance using clinical variables and concatenation of clinical variables with slide-level features from two multi-modal models (H&E + PR and All stains). All the models were optimised on the cohort subset (N=803), where the patients with missing clinical variables were excluded. In the clinical-only model, cox proportional hazard (PH) model was fitted on the well-established clinical variables (Age, Lymph node status, Tumour size, Histological grade) using 5-fold cross-validation (cv). The multi-modal models were re-optimised using 5-fold cv on the cohort subset for the direct comparison with the clinical-only model.

Lastly, we explored the assignment of tile-level attention weights from the ABMIL model to evaluate the extent to which the attention was placed on the same spatial regions of the tissue across the different modalities. We included individual stains (H&E-only and PR-only) and the concatenation of stains (H&E + PR) models for the analyses. Initially, wwe observed the pairwise Pearson correlation of the attention weights from tiles for each WSI in the three models (Figure 5a). The median slide-level correlation between attention weights for the H&E-only and PR-only models, and for the PR-only and H&E + PR models, were found to be close to zero. We observed a higher median correlation between attention weights for each WSI in the H&E-only and H&E + PR models (r = 0.32). Further, we observed the pairwise intersection of regions in high-attention tiles by considering the top 10%, 20% and 30% of the attention weights. Similarly, we observed a higher median intersection percentage between H&E-only and H&E+PR model in the top 10%, 20% and 30% of the attention weights (Figure 5b,c,d).

**Figure 5.**
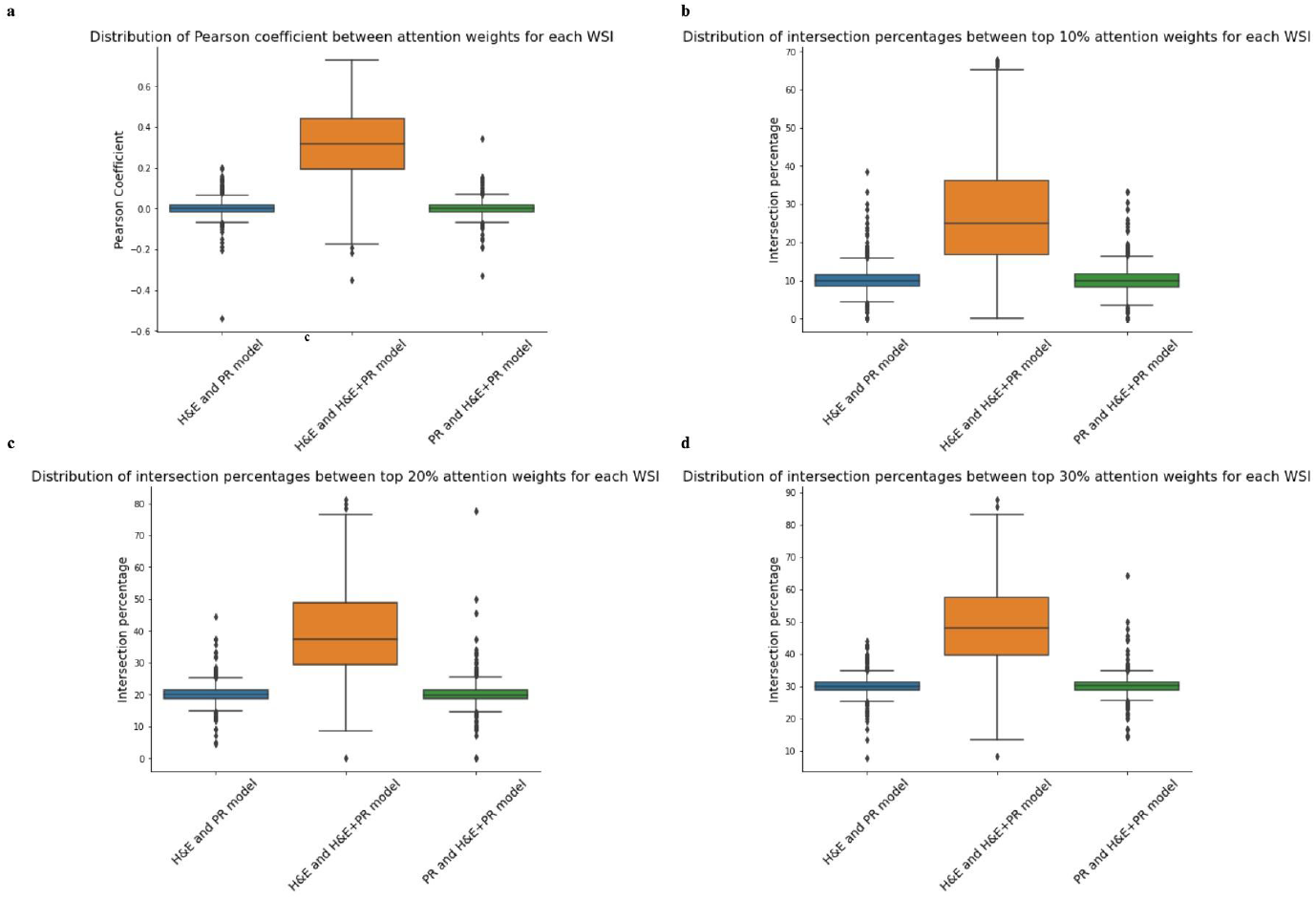
Evaluating the similarities in the assignment of attention weights to spatial regions across different prognostic models. Distribution analyses of tile-level attention weights for each whole-slide image (WSI) in three different models (H&E-only, PR-only and H&E+PR) was performed. **a** Distribution of pairwise Pearson correlation coefficient between tile-level attention weights for each WSI. **b,c,d** Distribution of intersection of percentages between top 10%, 20%, 30% tile-level attention weights for each WSI respectively.

## Discussion

In this study, we developed deep learning-based multi-stain model to predict prognostic risk score for breast cancer patients. We used the routine IHC WSIs, i.e. ER, PR, HER2 and Ki-67, along with H&E WSIs for the joint modelling for risk score prediction. H&E and IHC WSIs were obtained from the same tumour block. Further, we performed WSI registration between H&E and each IHC WSI to include more local combinations of histo-morphological information between different modalities.

We observed better risk score prediction performance using H&E-only compared to other individual stain modalities with both UNI and CONCH (C-index: 0.65 (95%CI: 0.57 - 0.74) and C-index: 0.64 (95%CI: 0.56 - 0.72)). Since H&E stained tissue is expected to capture a broader set of morphological information, compared to IHC, these results are not surprising. However, UNI had been pre-trained using only H&E WSIs, therefore extracting prognostic features from the IHC WSIs was not optimal but it still showed similar prognostic performance to CONCH that had been pre-trained using differently stained pathology images.

The addition of different stain modalities further improved the risk score prediction performance. The combination of H&E and PR WSIs provided similar prognostic performance (c-index: 0.72 (95% CI: 0.67 - 0.80)) in comparison to the combination of all stain modalities (c-index: 0.72 (95% CI: 0.66 - 0.81)) (Figure 3b).

Further, addition of the clinical variables to the slide-level features in the multi-modal models improved the predictive performance for the H&E + PR model but not for the All stain model (Figure 4). However, we observed better performance with multi-modal models when compared to the clinical-only model. It shows the combination of multi-stain modelling with known clinical variables can potentially improve the risk-stratification performance. The improvement by the addition of clinical variables in the H&E+PR model is quite apparent but the improvement was not observed in the all stain model when compared to the all stain model without the clinical variables. However, around 15% of the patients were excluded due to missingness in clinical variables which further decreased the statistical power with respect to survival outcome in this study.

Lastly, we explored the attention weights from the ABMIL model for individual and two stain models (H&E+PR). The spatial morphological features from the H&E WSI seem to contribute higher than the PR WSI, given the higher median correlation between the tile-level attention weights from the H&E-only and H&E+PR models in comparison to the PR-only and H&E+PR models (Figure 5a). It is further confirmed by observing similar patterns for the intersection percentages in the top 10, 20 and 30% percentage of the attention weights (Figure 5b,c,d). High-attention spatial regions from the H&E-only and PR-only models seem to be very different with median correlation close to zero, suggesting potentially independent prognostic information in these modalities with respect to the spatial localisation indicated by the attention weights. This further motivates exploration of more spatial interaction models between different stain modalities to improve prognosis prediction in future.

Previously, Song et al. [19] reported the 5-fold cv risk score prediction performance against the state of the art methods in multiple cancer types on the TCGA cohort. They used disease-specific survival as the survival endpoint to optimise the models. Focusing on breast cancer risk score prediction performance in TCGA-BRCA, the point estimates for H&E-based prognostic performance were observed to be similar (C-index: 0.67 and 0.65 for our model). When observing the multimodal prognostic performance estimates, we observed that the multi-stain modelling-based estimates were observed to be similar to the reported multimodal prototyping method (C-index: 0.75 and 0.72 for our model). However, both studies need external validation and given the differences in the cohorts and survival endpoints, a direct comparison is hard to establish.

The patient risk score prediction using routinely-stained WSI provides an advantage over existing molecular profiling-based methods by reducing the lead-times, cost and tissue damage among other factors [35]. However, one of the key limitations of this study is the absence of external validation sets to test the reproducibility and reliability of the models. The study also faces power issues with the number of events in the cohort leading to large spread in uncertainty estimates. Another limitation of this study is the relatively low median follow-up time (6.25 years) which increases the number of censored events in the cohort.

The future work includes the validation of models in larger independent external validation sets with longer follow-up time to establish the robustness and reliability of the multi-stain models that are established in this study. Further, we would like to explore more MIL-based methods that include interaction of feature vectors from different stain modalities. Some ideas like the Kronecker product of feature vectors other than the concatenation of feature vectors especially for two stain models can be utilised. Other MIL-based methods like TransMIL [36] involve more interaction between feature vectors than the ABMIL and can potentially capture more local and spatial interaction between morphological features from different stain modalities. Further,

To conclude, this study demonstrated that the addition of routinely assessed IHC WSIs to the standard H&E WSIs improved the risk-stratification performance in breast cancer patients. The local and spatial combination of histo-morphological features from H&E and IHC stains derived using publicly available foundation models like UNI and CONCH further improves the prognosis prediction in breast cancer patients especially over single-stain models. The multi-stain model shows promise to improve the prognosis prediction and can potentially further improve the risk-stratification of breast cancer patients.

## Data Availability

Data cannot be made public due to local legislation since it contains sensitive patient data.

## Notes

### Competing Interest Statement

JH has obtained speaker's honoraria or advisory board remunerations from Roche, Novartis, AstraZeneca, Pfizer, Eli Lilly, MSD and Gilead, and has received institutional research support from Cepheid, Novartis, Roche and AstraZeneca. MR and JH are co-founders and shareholders of Stratipath AB. All other authors have declared no conflicts of interest.

### Funding Statement

This work was supported by funding from the Swedish Research Council, Swedish Cancer Society, Karolinska Institutet, ERA PerMed (ERAPERMED2019-224-ABCAP), VINNOVA and SweLIFE (SwAIPP project), MedTechLabs, Swedish e-science Research Centre (SeRC) - eCPC, Stockholm Region, Stockholm Cancer Society and Swedish Breast Cancer Association.

### Author Declarations

The study has approval from the Swedish Ethical Review Authority (2017/2106-31, with amendments 2018/1462-32 and 2019-02336).

